# Artificially sweetened beverage intake and risk of liver-related adverse events in individuals with MASLD: A prospective UK Biobank cohort study

**DOI:** 10.64898/2026.07.04.26357265

**Authors:** Ning Xu, Jiaxi Lin, Lihe Liu, Shiqi Zhu, Rui Li, Jinzhou Zhu, Chunfang Xu

## Abstract

**Purpose:** Metabolic dysfunction-associated steatotic liver disease (MASLD) is a major cause of chronic liver disease and liver-related morbidity worldwide. Although dietary factors may influence MASLD progression, the long-term liver-specific implications of artificially sweetened beverage (ASB) intake remain unclear. We aimed to examine the association between ASB intake and the risk of liver-related adverse events and liver-related death among individuals with MASLD.

**Methods:** This prospective cohort study included 50,562 participants with MASLD from the UK Biobank. ASB intake was assessed using 24-hour dietary recalls and categorized as 0, >0–1, and >1 serving/day. Multivariable Cox proportional hazards models were used to estimate hazard ratios (HRs) and 95% confidence intervals (CIs) for liver-related adverse events and liver-related death. Restricted cubic spline models were used to assess dose-response patterns, and competing-risk analyses were performed by treating liver-related death as a competing event for liver-related adverse events. Additional substitution, subgroup and sensitivity analyses were conducted to evaluate the robustness of the findings.

**Results:** During a median follow-up of 12.8 years, 292 liver-related adverse events and 91 liver-related deaths occurred. Compared with participants reporting no ASB intake, those consuming >1 serving/day had a higher risk of liver-related adverse events in the fully adjusted model (HR 1.40, 95% CI 1.02–1.93; P = 0.039), whereas the association for >0–1 serving/day was not statistically significant (HR 1.26, 95% CI 0.92–1.71; P = 0.149). The risk of liver-related adverse events increased across ASB intake categories (P for trend = 0.023). Restricted cubic spline analysis indicated a positive linear association between ASB intake and liver-related adverse events (P-overall <0.001; P-nonlinearity = 0.72). In competing-risk analysis, the association for >1 serving/day remained consistent after accounting for liver-related death as a competing event (sub-HR 1.40, 95% CI 1.02–1.93; P = 0.038; Gray test P = 0.006). The association was robust in sensitivity analyses. ASB intake was not significantly associated with liver-related death, and beverage substitution analyses showed no significant associations.

**Conclusion:** Among individuals with MASLD, high ASB intake, particularly >1 serving/day, was associated with an increased risk of liver-related adverse events, but not liver-related death. This association was consistent across dose-response, competing-risk, and sensitivity analyses, suggesting that high ASB intake may represent a potential dietary risk marker for adverse liver outcomes in MASLD.

## Introduction

Metabolic dysfunction-associated steatotic liver disease (MASLD) is a chronic liver condition defined by hepatic steatosis in the presence of cardiometabolic risk factors. With the increasing prevalence of obesity, type 2 diabetes, dyslipidemia, and hypertension, MASLD has become one of the leading causes of chronic liver disease worldwide[1]. Although many patients remain clinically stable, a subset may progress to advanced fibrosis, cirrhosis, portal hypertension, liver failure, hepatocellular carcinoma, and liver-related death[2]. This growing clinical burden has led to increasing interest in modifiable factors that may influence MASLD progression, including dietary behaviors and beverage consumption patterns.

Dietary factors are closely linked to metabolic health and may contribute to liver fat accumulation, inflammation, fibrosis, and long-term liver outcomes[3]. Sugar-sweetened beverages have been associated with obesity, insulin resistance, type 2 diabetes, and fatty liver disease[4]. In response to concerns regarding sugar intake, artificially sweetened beverages (ASBs) are widely consumed as low-calorie alternatives, particularly among individuals with obesity, diabetes, or other metabolic disorders[5]. However, the absence of sugar does not necessarily indicate metabolic neutrality. Experimental and epidemiological evidence suggests that ASBs may affect glucose regulation, appetite control, gut microbiota composition, systemic inflammation, and cardiometabolic risk[6]. Nevertheless, the health effects of ASBs remain controversial, and most available studies have focused on metabolic traits or incident cardiometabolic diseases rather than liver-specific clinical outcomes.

For individuals with MASLD, this uncertainty is particularly important. These patients already have an adverse metabolic background and may be more susceptible to dietary exposures that influence metabolic regulation, the gut-liver axis, and hepatic disease progression[7]. Despite the widespread consumption of ASBs, few large-scale prospective studies have examined whether ASB intake is associated with clinically meaningful liver outcomes in MASLD, such as liver-related adverse events or liver-related death. It also remains unclear whether such associations show a dose-response pattern or persist after accounting for competing risks and potential sources of bias.

Therefore, using data from the UK Biobank, we conducted a prospective cohort study to evaluate the association between ASB intake and the risk of liver-related adverse events and liver-related death among individuals with MASLD. We further examined dose-response patterns and performed competing-risk, substitution, subgroup, and sensitivity analyses to assess the robustness and clinical relevance of the observed associations.

## Methods

### Design of the study and population

Our prospective study utilized data from the UK Biobank, a large population-based cohort covering the entire United Kingdom. Between 2006 and 2010, the UK Biobank recruited more than 500,000 people from 22 institutions across the UK[8]. The UK Biobank research protocol was approved by the South West Multicenter Research Ethics Committee. The study we present has been approved by the UK Biobank Scientific Committee. For more information on the UK Biobank research protocol, please click here (https://www.ukbiobank.ac.uk). Data are available on application. This research was conducted using the application number 373,472.

Among the 502,258 participants recruited by the UK Biobank between 2006 and 2010, we first excluded individuals with missing follow-up data, pre-existing liver-related adverse events and alcohol use disorder at baseline, including alcoholic liver disease (ALD), viral hepatitis, autoimmune liver disease, biliary cirrhosis, and primary liver cancer(Additional file1: Table S2), as well as those who did not meet the diagnostic criteria for MASLD, leaving 122,102 eligible participants. Participants without 24-hour dietary recall data, with unreliable dietary data, or with more than 10% missing covariate data were then further excluded. Ultimately, 50,562 individuals with MASLD were retained for the final analysis (Fig 1).

**Fig 1.**
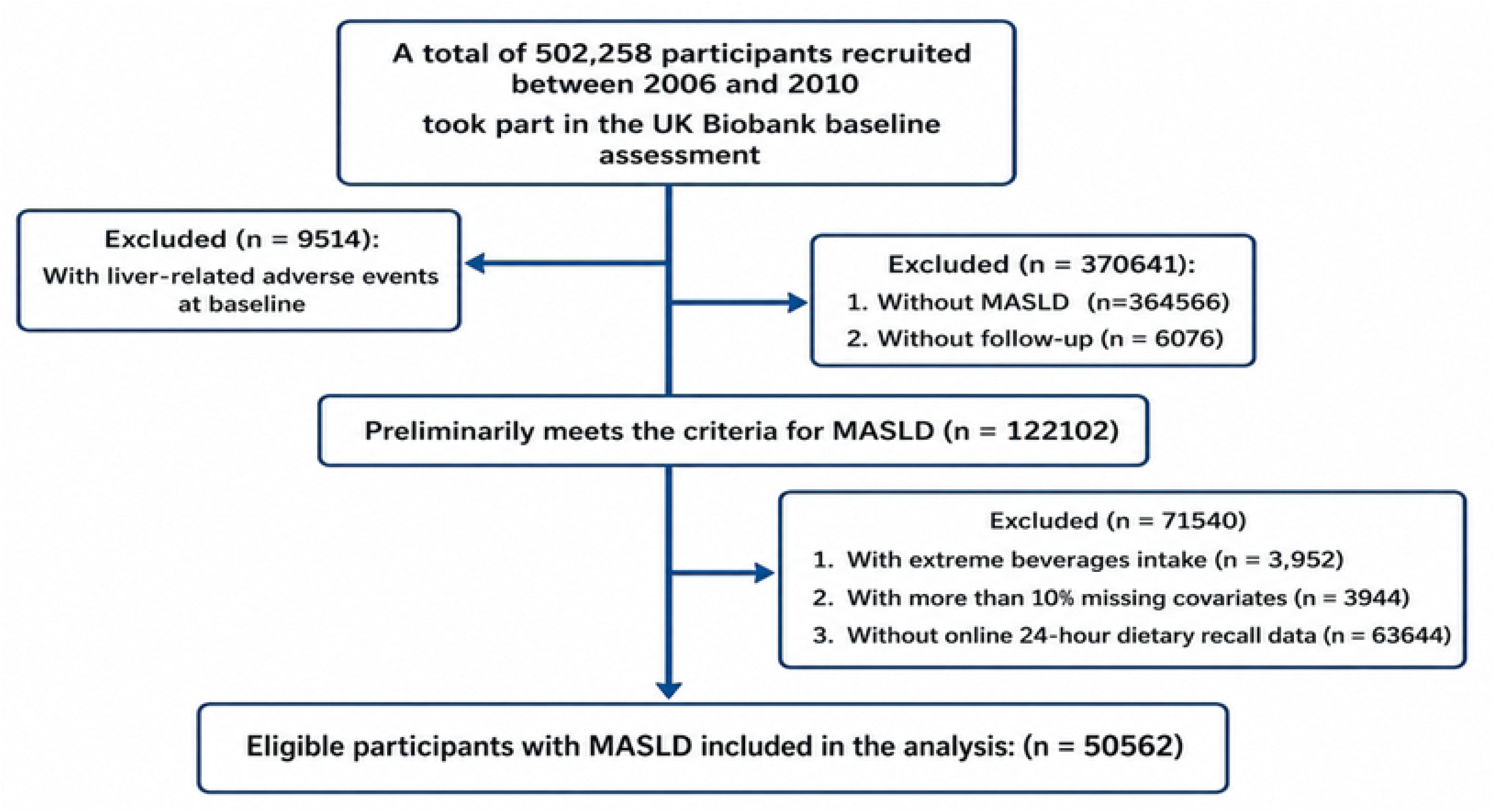
Flow chart of participant enrolment. Note: MASLD, Metabolic dysfunction-associated steatotic liver disease.

### Assessment of exposure

Dietary intake was assessed using the Oxford Web-Q, a web-based 24-hour dietary recall questionnaire. The questionnaire was administered up to five times between April 2009 and June 2012. Participants reported their intake of artificially sweetened beverages (ASBs) and natural juices (NJs) during the preceding 24 hours by selecting the number of servings consumed, defined as glasses, cans, or 250-mL cartons. Usual intake was estimated as the mean intake across available dietary recalls. Low-calorie beverages were classified as ASBs, whereas pure fruit and vegetable juices were classified as NJs. Participants were then categorized into three groups according to ASB intake: 0, >0–1, and >1 serving/day.

### Covariates

Covariates were selected based on previous evidence and their potential associations with beverage intake and liver-related outcomes. These covariates included demographic and socioeconomic factors, including age, sex, ethnicity, BMI category (< 25, ≥ 25 & < 30, and≥ 30 kg/m^2^), educational attainment, and Townsend deprivation index; lifestyle and dietary factors, including physical activity, total energy intake, total sugar intake, diet quality score, smoking status, and alcohol consumption; and clinical factors, including medication use(such as antihypertensive, antidiabetic, and lipid-lowering medications, etc.) and diabetes. The diet quality score (Additional file1: Table S12) was constructed according to the intake of processed meat, fish, vegetables, and fruits. Detailed definitions of all covariates are provided in the Supplementary Materials (Additional file1: Table S1).

### Definition of MASLD

MASLD was defined as the presence of hepatic steatosis together with at least one cardiometabolic risk factor[9]. In contrast to previous studies and European clinical practice guidelines that used a fatty liver index (FLI) ≥60 to define hepatic steatosis[10–12], the present study used a hepatic steatosis index (HSI) ≥36 as a feasible non-invasive surrogate when imaging-based assessment was unavailable. The HSI is a validated prediction index for hepatic steatosis based on the alanine aminotransferase/aspartate aminotransferase ratio, body mass index (BMI), sex, and diabetes status[13]. Compared with the FLI, the HSI is simpler to calculate, requires fewer variables, and does not rely on waist circumference or γ-glutamyl transferase, which may be frequently missing in large epidemiological databases. Therefore, the HSI was considered more operationally feasible for this large-scale cohort study[14]. The HSI was calculated as follows: HSI = 8 × (alanine aminotransferase/aspartate aminotransferase ratio) + BMI (+2 if female; +2 if diabetes). Diabetes was defined as described above.

Cardiometabolic risk factors were defined as follows: (1) blood pressure ≥130/85 mmHg or current use of antihypertensive medication; (2) fasting plasma glucose ≥100 mg/dL, glycated hemoglobin ≥6.5%, or a diagnosis of or treatment for type 2 diabetes; (3) BMI ≥25 kg/m² or waist circumference ≥94 cm in men or ≥80 cm in women; (4) triglycerides ≥150 mg/dL or current use of lipid-lowering medication; and (5) high-density lipoprotein cholesterol (HDL-C) <40 mg/dL in men or <50 mg/dL in women, or current use of lipid-lowering medication. Participants with hepatic steatosis and cardiometabolic dysfunction but alcohol intake exceeding the MASLD threshold were classified as having metabolic dysfunction-associated alcohol-related liver disease (MetALD) rather than MASLD. Therefore, MASLD was restricted to participants with an average alcohol intake of <30 g/day in men and <20 g/day in women.

### Ascertainment of outcome

The primary outcome was the occurrence of liver-related adverse events during follow-up. Liver-related adverse events were defined as a composite outcome based on ICD-10 codes, including hepatic fibrosis (K74.0), portal hypertension (K76.6), ascites (R18), liver failure (K72.0, K72.1, and K72.9), and other major liver-related complications. Liver-related death was evaluated as a secondary outcome and was defined as death for which liver disease was recorded as the primary or secondary cause on the death certificate. Outcomes were ascertained through linkage to hospital inpatient records, death registry data, and coded primary care records. For each participant, follow-up time was calculated from the date of baseline assessment to the first occurrence of a liver-related outcome, death, loss to follow-up, or the end of follow-up on October 31, 2022, whichever occurred first. The ICD-10 codes used to define all outcomes are provided in the Supplementary Materials (Additional file 1: Table S3).

### Statistical analysis

Participants were categorized according to beverage intake. Cox proportional hazards regression models were used to estimate hazard ratios (HRs) and 95% confidence intervals (CIs) for the associations of ASBs intake with liver-related adverse events and liver-related death. The proportional hazards assumption was assessed using Schoenfeld residuals. Multivariable models were adjusted for age, sex, and other potential confounders, including race/ethnicity, BMI category, educational attainment, Townsend deprivation index, physical activity, total energy intake, total sugar intake, diet quality score, smoking status, alcohol consumption, medication use, and diabetes status. These covariates were selected based on previous evidence and their potential associations with liver-related adverse events and liver-related death[15]. Missing covariate data were imputed using multiple imputation by chained equations. Restricted cubic spline models were used to evaluate potential dose-response associations between ASB intake and liver-related outcomes.

Substitution analyses were further performed to estimate the potential effect of replacing ASBs with NJs or plain water on the risk of liver-related adverse events. Detailed descriptions of covariate assessment and statistical methods, including sensitivity and subgroup analyses, are provided in the Supplementary Materials. All statistical analyses were performed using R software, version 4.5.1. Two-sided P values <0.05 were considered statistically significant.

## Results

### Baseline features of the research sample

A total of 50,562 participants with MASLD were included in the present analysis. Among them, 36,822 participants (72.8%) reported no intake of ASBs, 7,720 (15.3%) consumed >0–1 serving/day, and 6,020 (11.9%) consumed >1 serving/day. Baseline characteristics according to ASBs intake are shown in Table 1.

**Table 1.**
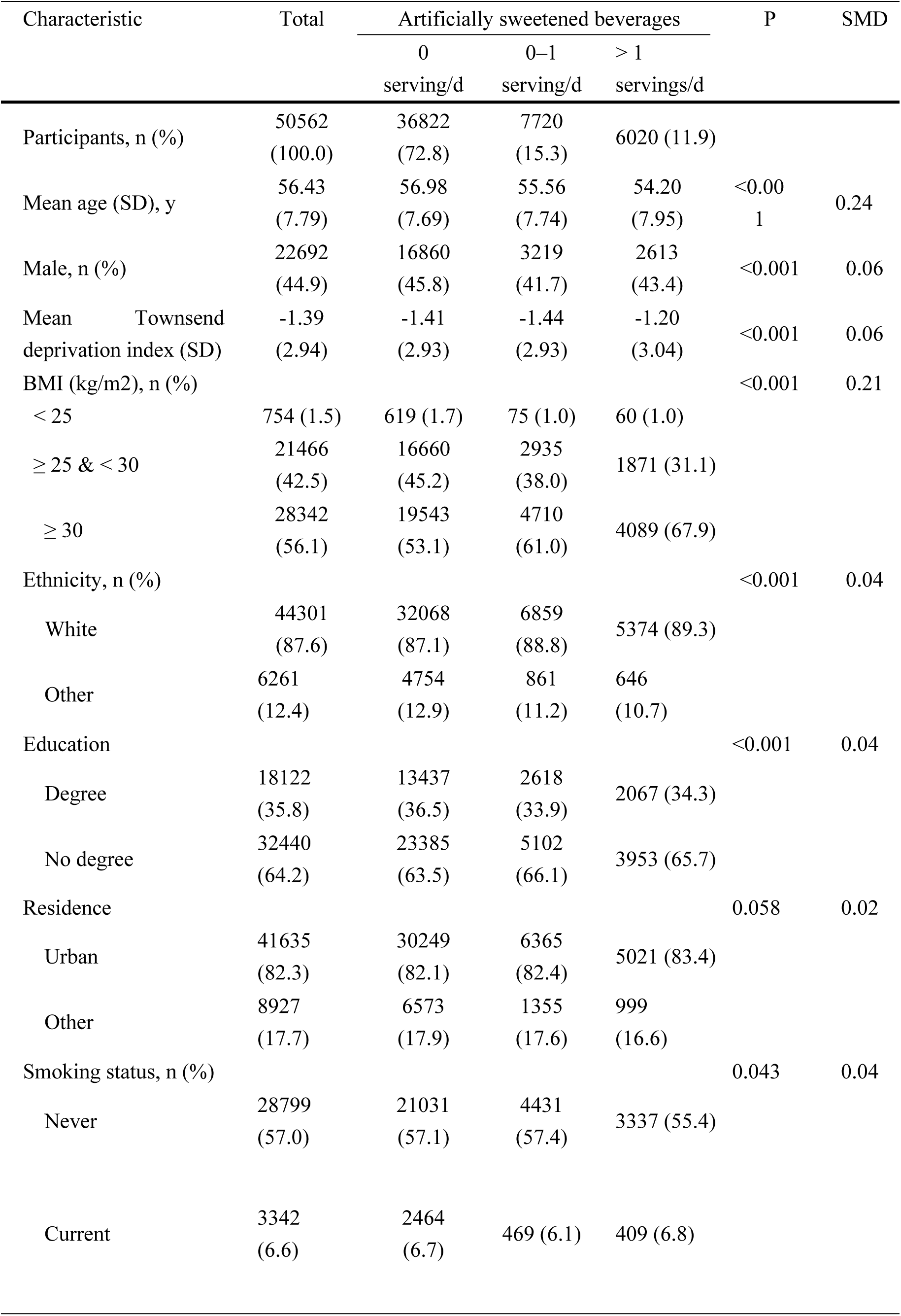
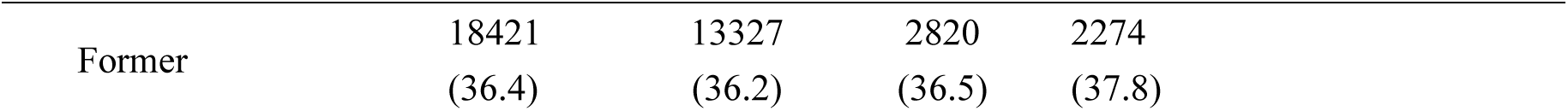
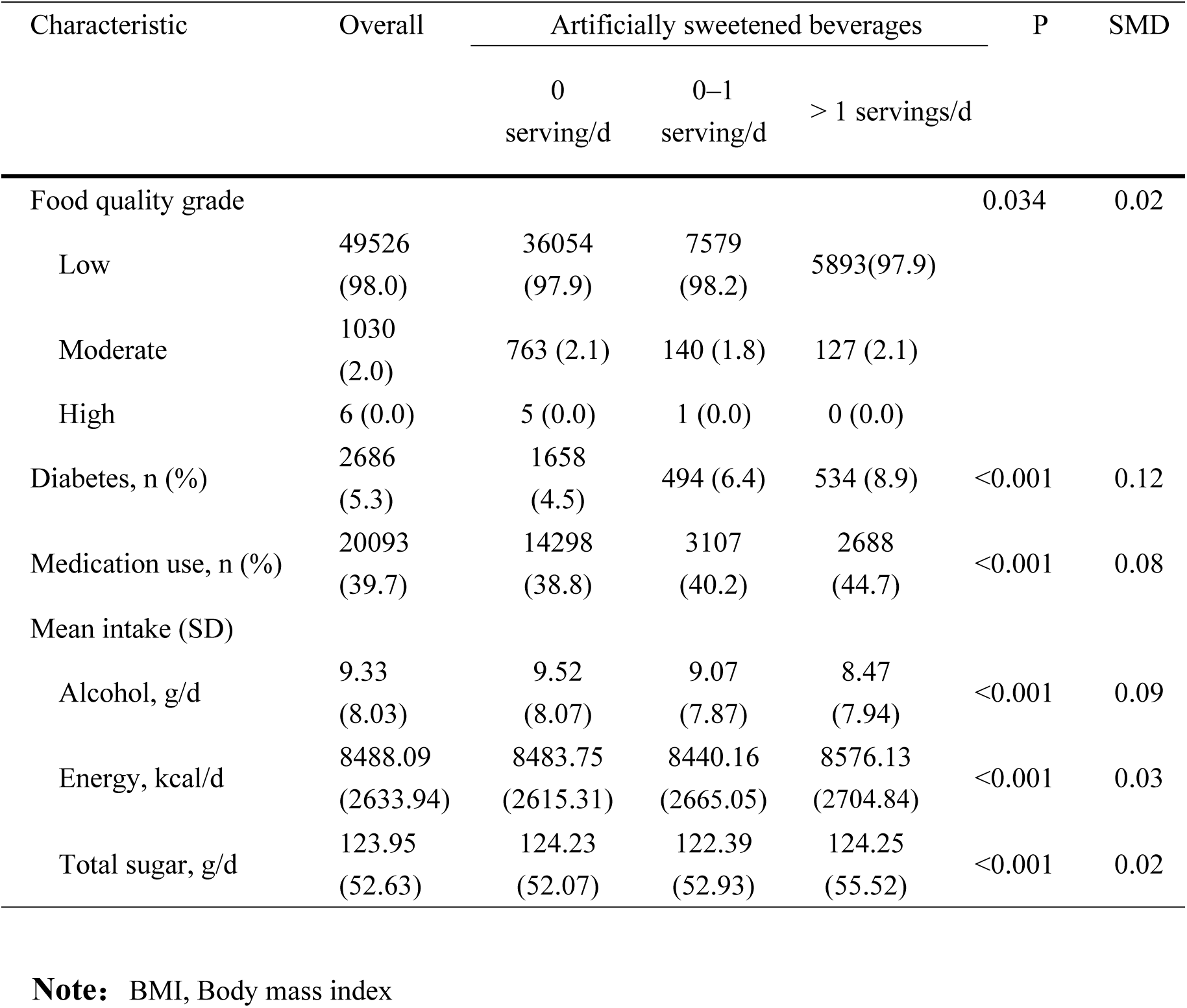
Basic characteristics of participants (ASBs)

Participants with higher ASBs intake differed from nonconsumers in several clinically relevant characteristics. Compared with nonconsumers, those consuming >1 serving/day of ASBs were younger [54.20 (7.95) vs. 56.98 (7.69) years] and had a less favorable metabolic profile, including a higher proportion of obesity defined as BMI ≥30 kg/m² [4,089 (67.9%) vs. 19,543 (53.1%)] and a higher prevalence of diabetes [534 (8.9%) vs. 1,658 (4.5%)]. They were also more likely to report medication use [2,688 (44.7%) vs. 14,298 (38.8%)]. Standardized mean differences suggested that the most notable baseline differences across ASBs intake categories were observed for age, BMI category, and diabetes status. Baseline characteristics according to NJs intake are presented in the Supplementary Materials (Additional file 1: Table S11).

### ASBs intake and liver-related adverse events

During follow-up (median: 12.8years, IQR: 12.2–13.6 years), 292 liver-related adverse events were documented, including 190 events among participants reporting no ASBs intake, 52 events among those consuming >0–1 serving/day, and 50 events among those consuming >1 serving/day.

Higher ASBs intake was associated with an increased risk of liver-related adverse events. Although the association was attenuated after multivariable adjustment, participants consuming >1 serving/day of ASBs consistently had a higher risk than nonconsumers across all models (Fig 2).

**Fig 2.**
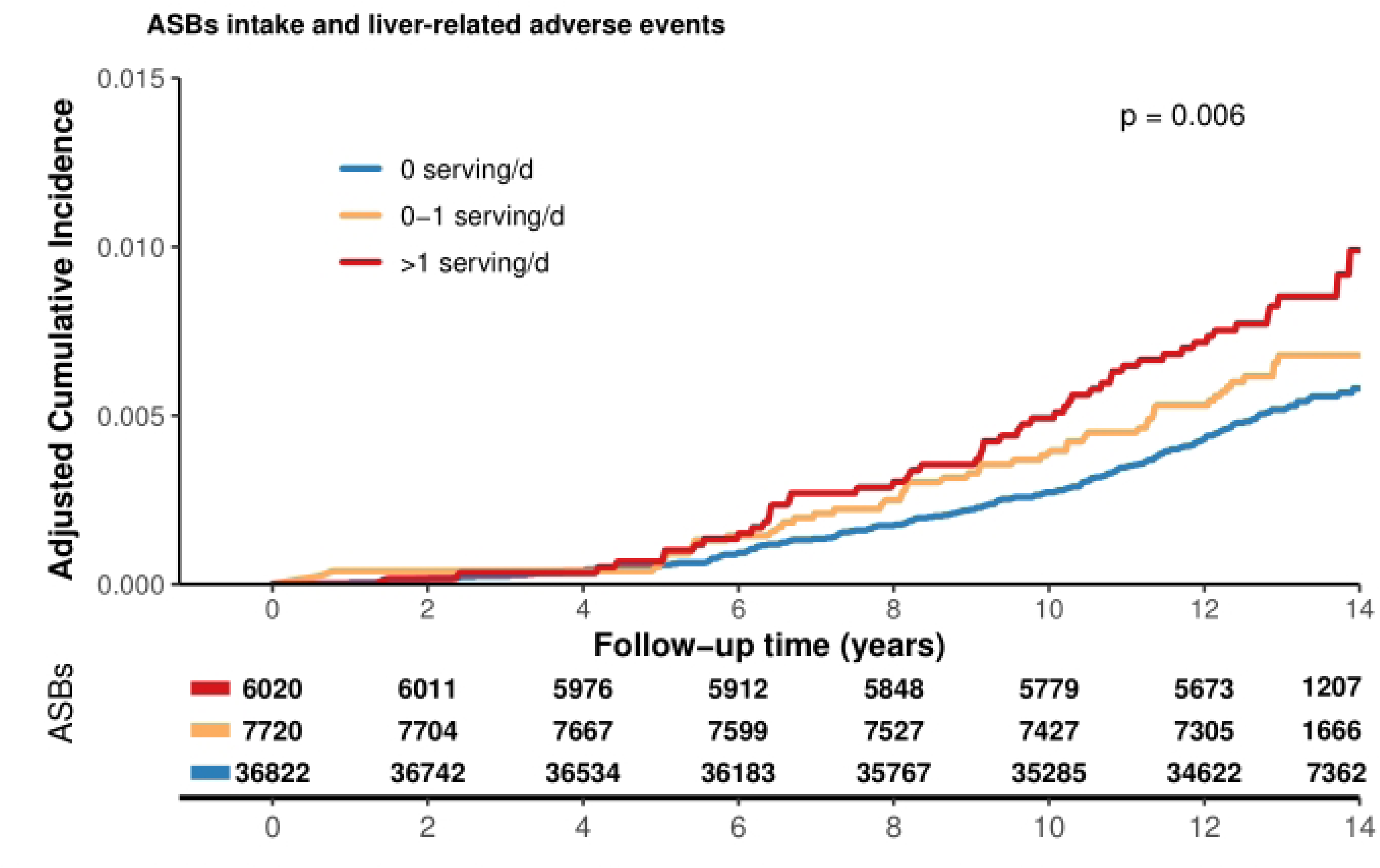
Multivariable-adjusted cumulative incidence of liver-related adverse events according to consumption of ASBs. Note: The model adjusted for the following variables: Age (continuous variable), Sex (male or female), BMI (< 25, ≥ 25, < 30, and ≥ 30 kg/m²), ethnicity (White or other), education level(degree or no degree), smoking status (current, former, or never), residence (urban or other), Townsend Deprivation Index (continuous), IPAQ activity group (low, moderate, high), alcohol consumption frequency (Never, Daily or almost daily, Three or four times a week, Once or twice a week, One to three times a month, Special occasions only), Food quality grade (Low, Moderate, High),daily alcohol intake (continuous variable), total energy intake(continuous), total sugar intake (continuous), medication use (yes or no), and diabetes (yes or no); and adjustments were made for natural juice and water. ASB,artificially sweetened beverage;BMI, Body mass index; MASLD, Metabolic dysfunction-associated steatotic liver disease. IPAQ, International Physical Activity Questionnaire.

In the fully adjusted model, compared with participants reporting no ASBs intake, those consuming >0–1 serving/day had a non-significantly higher risk of liver-related adverse events (HR 1.26, 95% CI 0.92–1.71; P = 0.149), whereas those consuming >1 serving/day had a significantly higher risk (HR 1.40, 95% CI 1.02–1.93; P = 0.039, Table 2). A significant trend was observed across increasing ASBs intake categories (P for trend = 0.023), suggesting a graded association between ASBs intake and liver-related adverse events.

**Table 2.** Associations between consumption of ASBs and risk of liver-related adverse events.

|  | Nonconsumers |  | ASB consumers |  |  |  | P for trend |
| --- | --- | --- | --- | --- | --- | --- | --- |
|  | 0 serving/d | P | 0–1 serving/d | P | >1 serving/d | P |  |
| Number of participants | 36822 |  | 7720 |  | 6020 |  |  |
| Number of liver-related adverse events | 190 |  | 52 |  | 50 |  |  |
| Model 1 | 1 (ref) | NA | 1.44<br>(1.06–1.96) | 0.020 | 1.89<br>(1.38–2.58) | <0.001 | <0.001 |
| Model 2 | 1 (ref) | NA | 1.36<br>(1.00–1.85) | 0.052 | 1.68<br>(1.22–2.30) | 0.001 | <0.001 |
| Model 3 | 1 (ref) | NA | 1.26<br>(0.92–1.71) | 0.149 | 1.40<br>(1.02–1.93) | 0.039 | 0.023 |

**Note:** Estimates are hazard ratios (95% confidence intervals), obtained from Cox regression models.
Model 1 was adjusted for age (continuous) and sex (male or female) and mutually adjusted for natural juice and water.
Model 2 was further adjusted for body mass index (< 25, ≥ 25 & < 30, and ≥ 30 kg/m<sup>2</sup>), ethnicity (White or other), Townsend deprivation index (continuous), education level (degree or no degree), smoking status (current, former, or never), residence (urban or other), IPAQ activity group (low, moderate, high).
Model 3 was further adjusted for intake of total energy (continuous), total sugar (continuous), alcohol consumption frequency (Never, Daily or almost daily, Three or four times a week, Once or twice a week, One to three times a month, Special occasions only), Food quality grade (Low, Moderate, High), daily alcohol intake (continuous variable), medication use (yes or no), and diabetes (yes or no). BMI, Body mass index; MASLD, Metabolic dysfunction-associated steatotic liver disease. IPAQ, International Physical Activity Questionnaire.
P for trend was calculated by using median value of ASBs intake (0, 0–1, and > 1) groups.

Restricted cubic spline analysis using ASBs intake as a continuous variable further supported this association. The overall association between ASBs intake and liver-related adverse events was statistically significant (P-overall <0.001), with no evidence of non-linearity (P-nonlinearity = 0.72, Fig 3), indicating an approximately linear increase in risk with higher ASBs intake. The proportional hazards assumption was not violated for either liver-related adverse events or liver-related death based on Schoenfeld residual tests (Additional file 1: Fig S1, Table S4). No significant association was observed between ASBs intake and liver-related death in eithercategorical or trend analyses.

**Fig 3.**
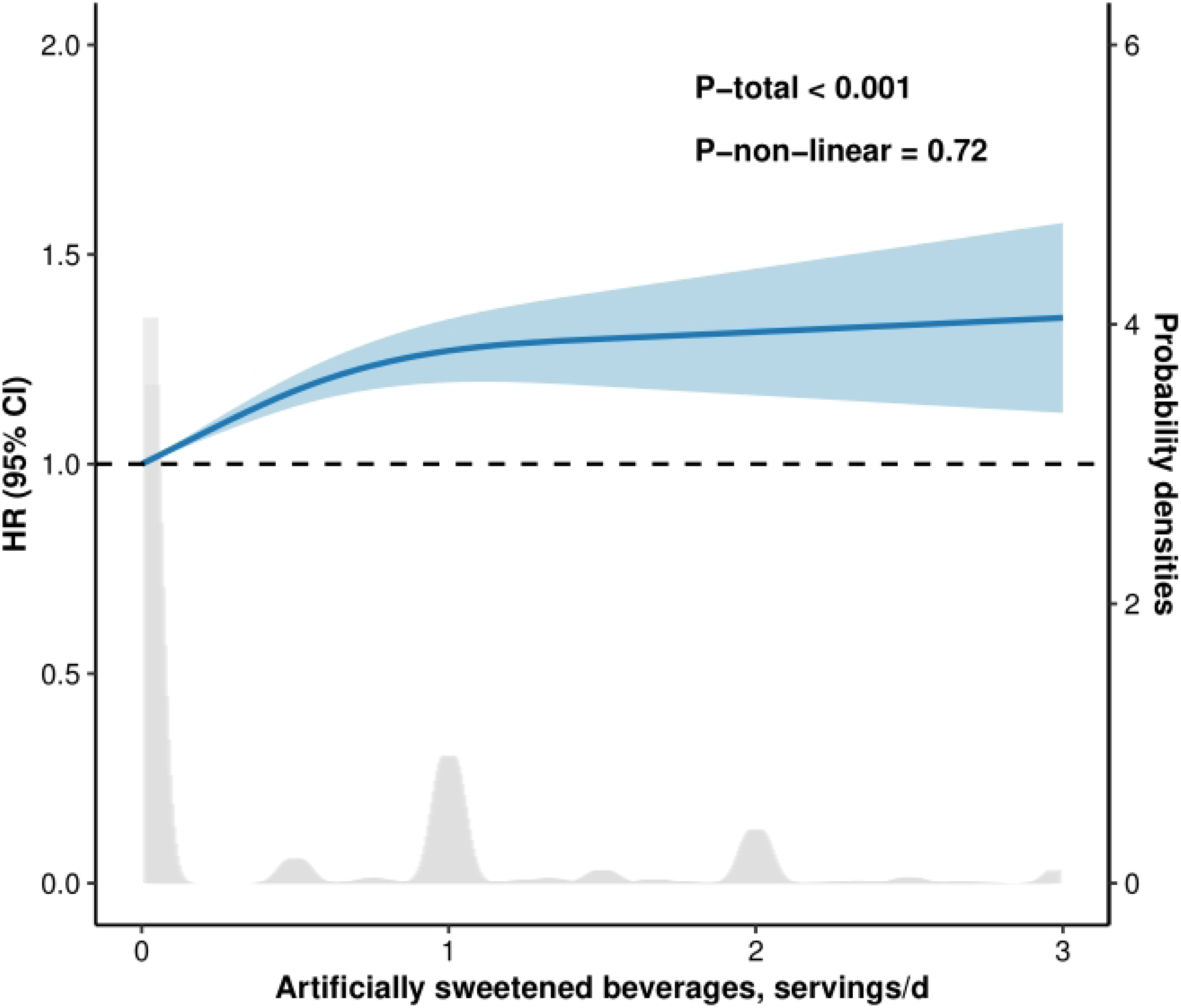
Restrictive cubic spline model of the association between intake of ASBs and the risk of liver-related adverse events during follow-up. Note: The model adjusted for the following variables: Age (continuous variable), Sex (male or female), BMI (< 25, ≥ 25, < 30, and ≥ 30 kg/m²), ethnicity (White or other), education level(degree or no degree), smoking status (current, former, or never), residence (urban or other), Townsend Deprivation Index (continuous), IPAQ activity group (low, moderate, high), alcohol consumption frequency (Never, Daily or almost daily, Three or four times a week, Once or twice a week, One to three times a month, Special occasions only), Food quality grade (Low, Moderate, High),daily alcohol intake (continuous variable), total energy intake(continuous), total sugar intake (continuous), medication use (yes or no), and diabetes (yes or no); and adjustments were made for natural juice and water. BMI, Body mass index; MASLD, Metabolic dysfunction-associated steatotic liver disease. IPAQ, International Physical Activity Questionnaire.

### Competing-risk analysis

In the competing-risk analysis, liver-related death was treated as a competing event for liver-related adverse events. The association between higher ASBs intake and liver-related adverse events remained consistent with the primary Cox analysis. Compared with participants reporting no ASBs intake, the subdistribution hazard ratio for liver-related adverse events was 1.26 (95% CI 0.92–1.72; P = 0.151) among those consuming >0–1 serving/day and 1.40 (95% CI 1.02–1.93; P = 0.038) among those consuming >1 serving/day. Cumulative incidence function curves showed progressive separation across ASBs intake categories, with the highest cumulative incidence observed among participants consuming >1 serving/day of ASBs (Gray test P = 0.006, Additional file 1: Fig S2). These findings suggest that the positive association between high ASB intake and liver-related adverse events was not materially altered after accounting for liver-related death as a competing event. Results for liver-related death in the competing-risk framework were not statistically significant (Additional file 1: Table S5).

### Beverage substitution analysis

Model-based substitution analyses were performed to explore whether replacing ASBs intake with alternative beverages was associated with liver-related outcomes. In the fully adjusted model, replacing one serving/day of ASBs with water was not significantly associated with the risk of liver-related adverse events (HR 0.94, 95% CI 0.82–1.07; P = 0.321). Similarly, replacing one serving/day of ASBs with natural juices was not significantly associated with liver-related adverse events (HR 0.97, 95% CI 0.78–1.21; P = 0.808). Similar null findings were observed when total energy intake and total sugar intake were excluded from the substitution models.Parallel substitution analyses for liver-related death also showed no significant associations (Additional file 1: Table S6).

### Subgroup and sensitivity analyses

In subgroup analyses for liver-related adverse events, the positive association between high ASBs intake and liver-related adverse events was generally consistent across major demographic, metabolic, and lifestyle subgroups. No significant interactions were observed for sex, age, diabetes status, medication use, food quality, or alcohol intake. A significant interaction was observed for BMI category (P for interaction = 0.003). However, subgroup-specific estimates among participants with BMI <25 kg/m² were based on very few events, resulting in unstable estimates and wide or non-estimable confidence intervals. Therefore, the BMI-specific findings should be interpreted cautiously and considered exploratory. Subgroup analyses for liver-related death did not show significant effect modification (Additional file 1: Fig S3).

The main association between high ASBs intake and liver-related adverse events remained robust in sensitivity analyses. After excluding liver-related adverse events that occurred within the first year of follow-up, participants consuming >1 serving/day of ASBs continued to have a higher risk of liver-related adverse events compared with those reporting no ASBs intake (HR 1.42, 95% CI 1.03–1.95; P = 0.032). The association for >0–1 serving/day remained non-significant (HR 1.20, 95% CI 0.87–1.64; P = 0.265).When total energy intake and total sugar intake were removed from the fully adjusted model, the association between >1 serving/day of ASB intake and liver-related adverse events was essentially unchanged (HR 1.40, 95% CI 1.02–1.92; P = 0.040), whereas the association for >0–1 serving/day remained non-significant (HR 1.26, 95% CI 0.92–1.71; P = 0.148). Substitution analyses excluding total energy and sugar intake also showed no significant associations when ASBs were replaced with water or natural juices. The E-value for the association between >1 serving/day of ASB intake and liver-related adverse events was 2.15, with an E-value of 1.15 for the lower confidence limit, suggesting that an unmeasured confounder of moderate strength would be required to fully explain the observed association(Additional file 1: Table S7-S10).

## Discussion

In this large prospective cohort study of 50,562 individuals with MASLD from the UK Biobank, we found that higher intake of ASBs was associated with an increased risk of liver-related adverse events. Participants consuming more than one serving of ASBs per day had a significantly higher risk of liver-related adverse events than those reporting no ASBs intake, whereas the association for moderate ASBs intake (>0–1 serving/day) did not reach statistical significance. The positive association was supported by restricted cubic spline analysis, which suggested an overall linear increase in risk with increasing ASBs intake. Furthermore, the association persisted in competing-risk analysis after accounting for liver-related death as a competing event, indicating that the observed relationship was not materially explained by competing mortality.

Our findings add to the growing literature on the potential health consequences of ASBs consumption. ASBs are commonly consumed as low-calorie alternatives to sugar-sweetened beverages, particularly among individuals with obesity, diabetes, or other metabolic disorders[16]. However, increasing evidence suggests that the absence of sugar does not necessarily imply metabolic neutrality[17]. Previous studies have linked ASBs intake to adverse cardiometabolic outcomes, including weight gain, type 2 diabetes, cardiovascular disease, and metabolic dysfunction[18–20].

Nevertheless, evidence regarding liver-specific outcomes has been limited, especially in individuals with established MASLD. By focusing on liver-related adverse events rather than metabolic traits alone, the present study extends prior evidence and suggests that high ASB intake may be a marker of increased risk for clinically relevant liver disease progression in MASLD.

Several biological and behavioral pathways may help explain the observed association. First, ASBs consumption may reflect an underlying unfavorable metabolic profile[21,22]. In our study, participants with higher ASBs intake had a higher prevalence of obesity, diabetes, and medication use at baseline, suggesting that ASBs consumers may already represent a metabolically vulnerable subgroup. Although we adjusted for major demographic, lifestyle, dietary, and metabolic factors, residual confounding and confounding by indication cannot be fully excluded. Second, artificial sweeteners may influence metabolic regulation through effects on glucose homeostasis, insulin response, appetite control, and energy balance[23,24]. These changes may be particularly relevant in MASLD, a condition closely linked to insulin resistance and systemic metabolic dysfunction. Third, ASBs may affect the gut-liver axis[25,26]. Experimental and epidemiological evidence has suggested that certain artificial sweeteners may alter gut microbiota composition, intestinal permeability, bile acid metabolism, and inflammatory signaling, all of which may contribute to liver inflammation, fibrogenesis, and disease progression. Finally, ASBs intake may cluster with other unmeasured dietary or behavioral patterns that increase liver-related risk[27,28].

The association between ASBs intake and liver-related adverse events was robust across several additional analyses. After excluding events occurring within the first year of follow-up, the association between high ASBs intake and liver-related adverse events remained significant, reducing the likelihood that the results were entirely driven by reverse causation. Similarly, removing total energy intake and total sugar intake from the fully adjusted model did not materially change the risk estimate, suggesting that the main association was not dependent on adjustment for these dietary factors. The E-value for the main association was 2.15, indicating that an unmeasured confounder would need to be associated with both high ASBs intake and liver-related adverse events by a risk ratio of approximately 2.15-fold, beyond the measured covariates, to fully explain the observed association. However, the E-value for the lower confidence limit was modest, and residual confounding remains possible.

In contrast, ASBs intake was not significantly associated with liver-related death. This finding should be interpreted cautiously because only 91 liver-related deaths occurred during follow-up, limiting statistical power and resulting in relatively imprecise estimates. Therefore, the absence of a statistically significant association with liver-related death should not be interpreted as evidence that ASBs intake has no effect on liver-related mortality. Rather, liver-related death should be regarded as an exploratory secondary outcome in the present analysis. Larger studies with more liver-related death events or longer follow-up are needed to clarify this relationship.

The beverage substitution analyses did not show significant associations when ASBs intake was replaced by water or natural juices. These findings suggest that, within the present data, there was insufficient evidence to conclude that model-based replacement of ASBs with water or natural juices was associated with a lower risk of liver-related adverse events or liver-related death. Several explanations are possible. First, substitution analyses are based on statistical modeling and do not directly reflect randomized beverage replacement interventions. Second, dietary intake was assessed using 24-hour dietary recalls, which may not fully capture long-term habitual intake or changes in beverage consumption over time[29,30]. Third, the health impact of replacing ASBs may depend on the type, amount, and context of the substitute beverage. Natural juices, for example, may contain vitamins and bioactive compounds, but they also contain natural sugars and may vary substantially in composition[31]. Therefore, the null substitution findings should be interpreted as exploratory rather than definitive evidence against potential benefits of beverage replacement[32].

A significant interaction was observed between BMI category and ASBs intake in relation to liver-related adverse events. However, this finding should be considered exploratory because some BMI subgroups, particularly participants with BMI <25 kg/m², had very few events, leading to unstable estimates and wide or non-estimable confidence intervals. The interaction may reflect differences in baseline metabolic risk, body fat distribution, insulin resistance, or patterns of ASBs consumption across BMI categories[33,34]. Alternatively, it may have arisen from sparse data in certain subgroups. Further studies using alternative adiposity measures, such as waist circumference, visceral adiposity, or body composition, are needed to determine whether obesity modifies the association between ASB intake and liver-related outcomes in MASLD.

This study has several strengths. First, it was based on a large prospective cohort of individuals with MASLD, allowing temporal assessment of ASBs intake before the occurrence of liver-related outcomes. Second, we focused on clinically meaningful liver-related adverse events, rather than only intermediate metabolic or biochemical markers. Third, we used multiple analytical approaches, including multivariable Cox models, restricted cubic spline analysis, competing-risk analysis, subgroup analysis, sensitivity analysis, substitution analysis, and E-value estimation, which together provided a comprehensive assessment of the association and its robustness. Fourth, linkage to hospital inpatient records, death registry data, and coded primary care records improved outcome ascertainment over long-term follow-up.

Several limitations should also be acknowledged. First, because this was an observational study, causality cannot be established. Although we adjusted for a broad range of demographic, socioeconomic, lifestyle, dietary, and clinical factors, residual confounding remains possible. In particular, participants with higher ASB intake had a less favorable metabolic profile at baseline, including higher BMI, more diabetes, and more frequent medication use, raising the possibility of confounding by indication. Second, ASB intake was assessed using 24-hour dietary recalls. Although repeated recalls were used when available, dietary measurement error and within-person variation over time could not be avoided. Third, hepatic steatosis was defined using the hepatic steatosis index (HSI) rather than imaging or histology, which may have introduced misclassification. Fourth, liver-related death was uncommon, limiting statistical power for this secondary outcome. Fifth, substitution analyses were model-based and should not be interpreted as equivalent to randomized beverage replacement interventions. Finally, the UK Biobank population is predominantly middle-aged and White, and participants may be healthier than the general population; therefore, the generalizability of these findings to other populations with different ethnic backgrounds, dietary patterns, and MASLD severity requires further investigation.

## Conclusion

In conclusion, this large prospective cohort study showed that higher ASBs intake, particularly >1 serving/day, was associated with an increased risk of liver-related adverse events among individuals with MASLD. This association was consistent across dose-response, competing-risk, and sensitivity analyses, but was not observed for liver-related death or in beverage substitution analyses.

By focusing on long-term liver-specific outcomes rather than metabolic traits alone, these findings highlight the potential relevance of ASBs consumption in dietary risk assessment for MASLD. Given the observational design, causality cannot be established, and high ASB intake should be interpreted as a potential dietary risk marker rather than a proven causal factor. Further mechanistic and interventional studies are needed to confirm these findings and determine whether reducing high ASBs intake can improve liver-related outcomes in MASLD.

## Data Availability

For more information on the UK Biobank research protocol, please click here (https://www.ukbiobank.ac.uk). Data are available on application. This research was conducted using the application number 373,472.

## Supporting information

**Additional file 1.** The Additional file contains the variable IDs, the ICD-10 codes for the outcomes, and the necessary data analysis results.(DOCX)

## Acknowledgements

This research was conducted by using the UK Biobank resource under Application Number 373472. The authors would like to thank the UK Biobank participants and investigators for making this study possible. The authors highly appreciated the support from AIMprism team.

## Author contributions

NX and JL : conception and design, data analysis, and drafted the manuscript.

SZ and LL:data acquisition and data curation

RL:data preprocessing and visualization

CX and JZ: study supervision, revised the manuscript, and approved the final version.

All authors have read and approved the final manuscript.

## Supporting information

Table S1. Information about all variables involved in this study

Table S2. ICD-10 Codes for exclusion criteria

Table S3. ICD-10 Codes for Defining Outcomes

Table S4. Schoenfeld residual tests

Table S5. Competing-risk analysis

Table S6. Beverage substitution analysis

Table S7-10. Sensitivity analysis

Table S11. Basic characteristics of participants(NJs)

Table S12. Healthy diet score

**Figure S1.**
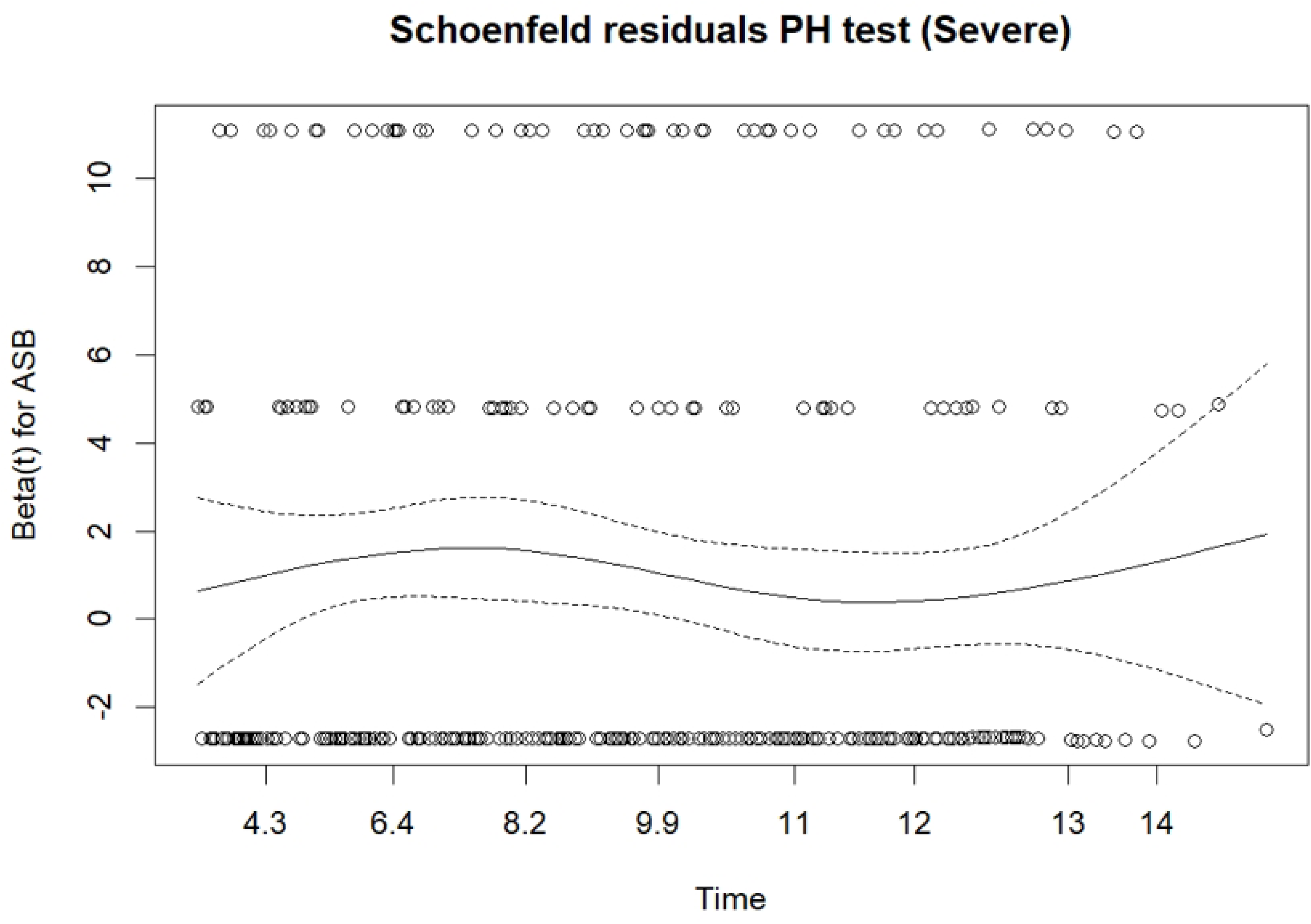
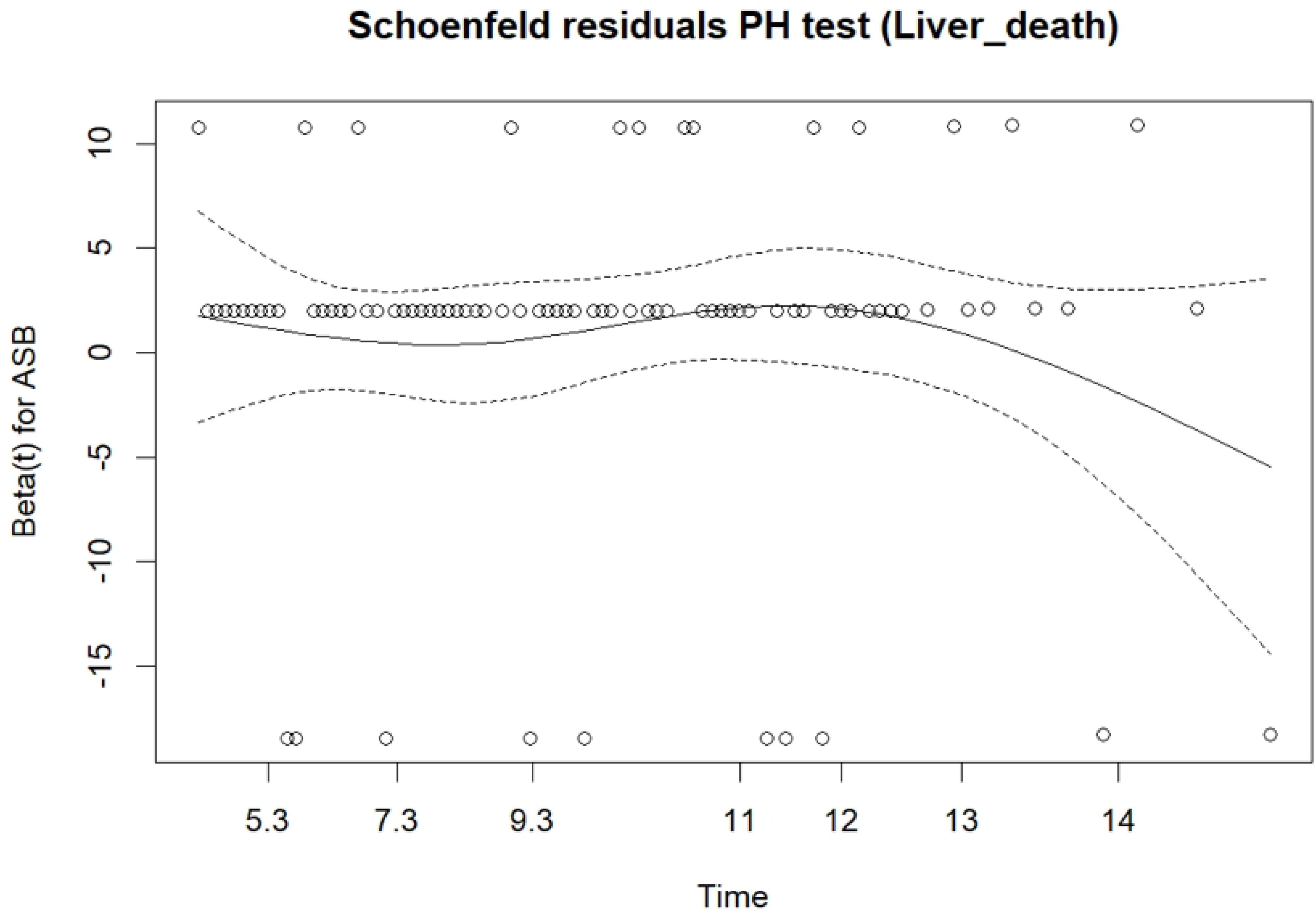
Schoenfeld residuals plot for proportional hazards assumption test of ASBs intake in association with liver-related adverse events and liver-related death

**Figure S2.**
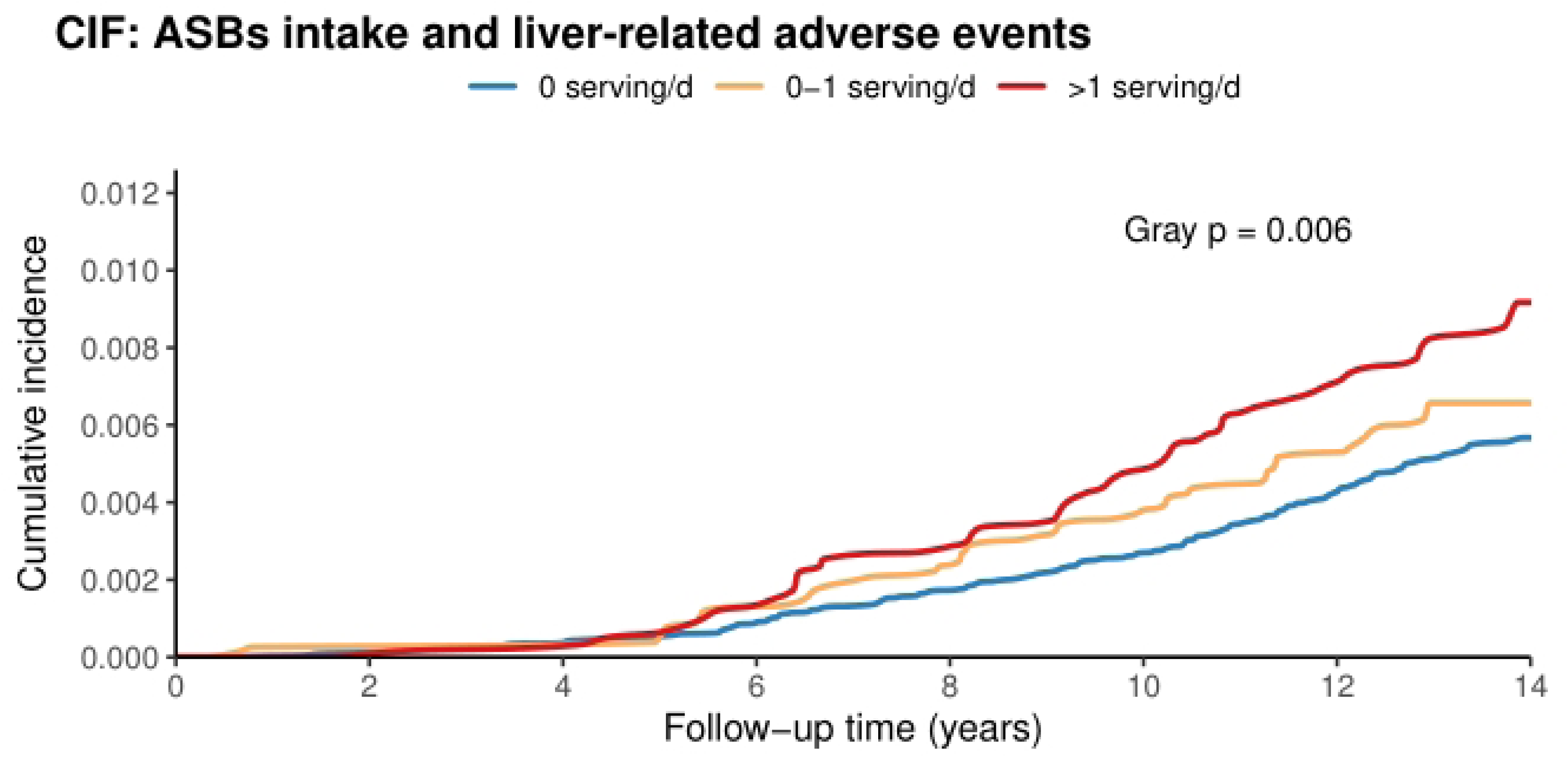
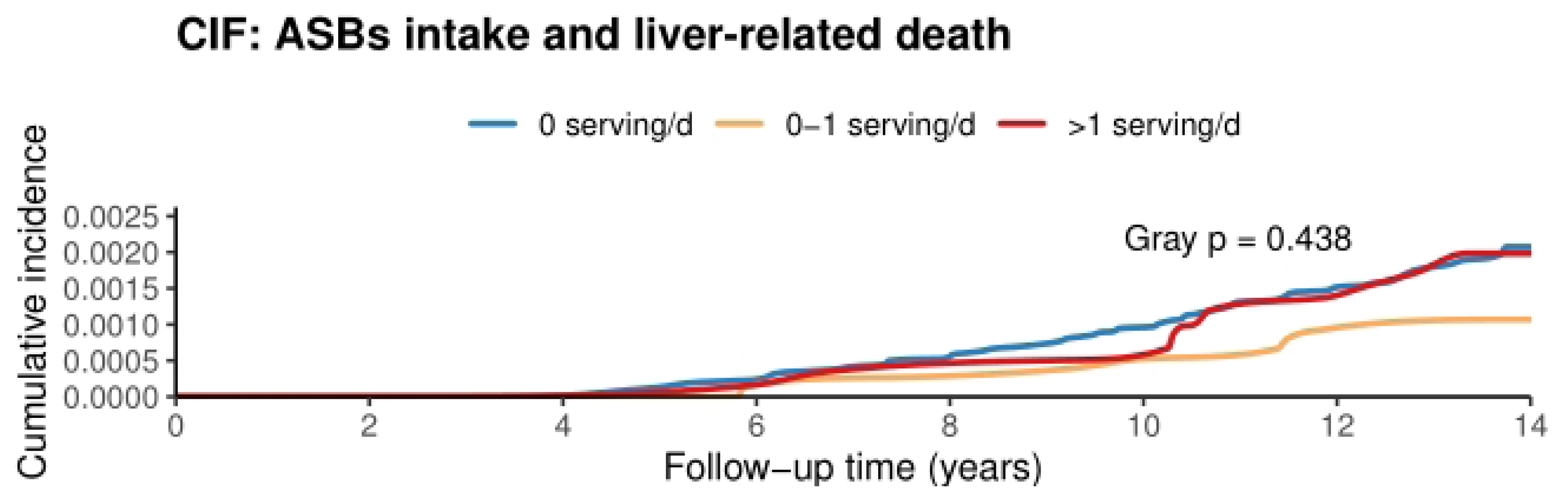
Cumulative incidence function curves of liver-related adverse events/ liver-related death across categories of ASBs intake accounting for competing risks

**Figure S3.**
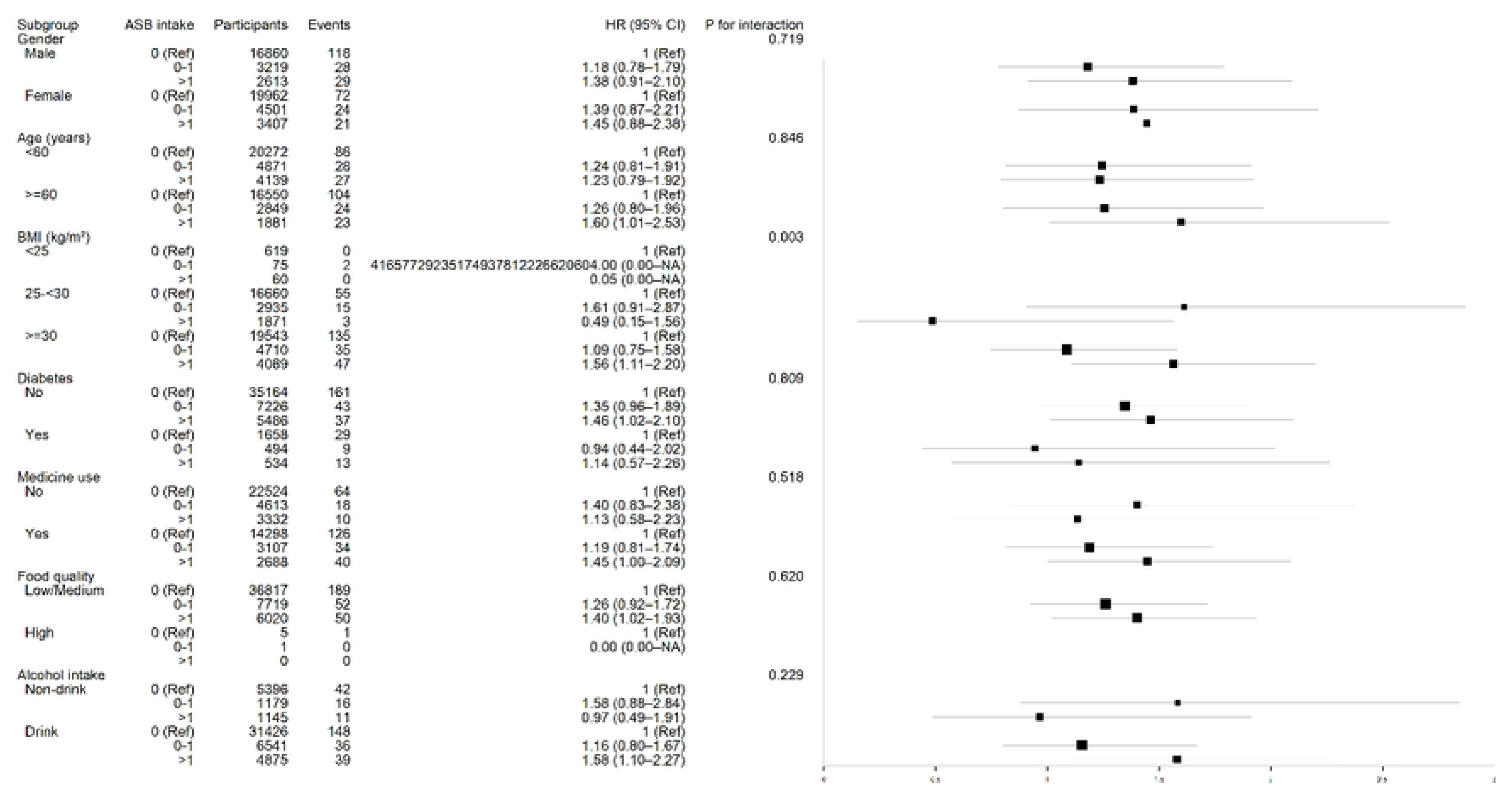
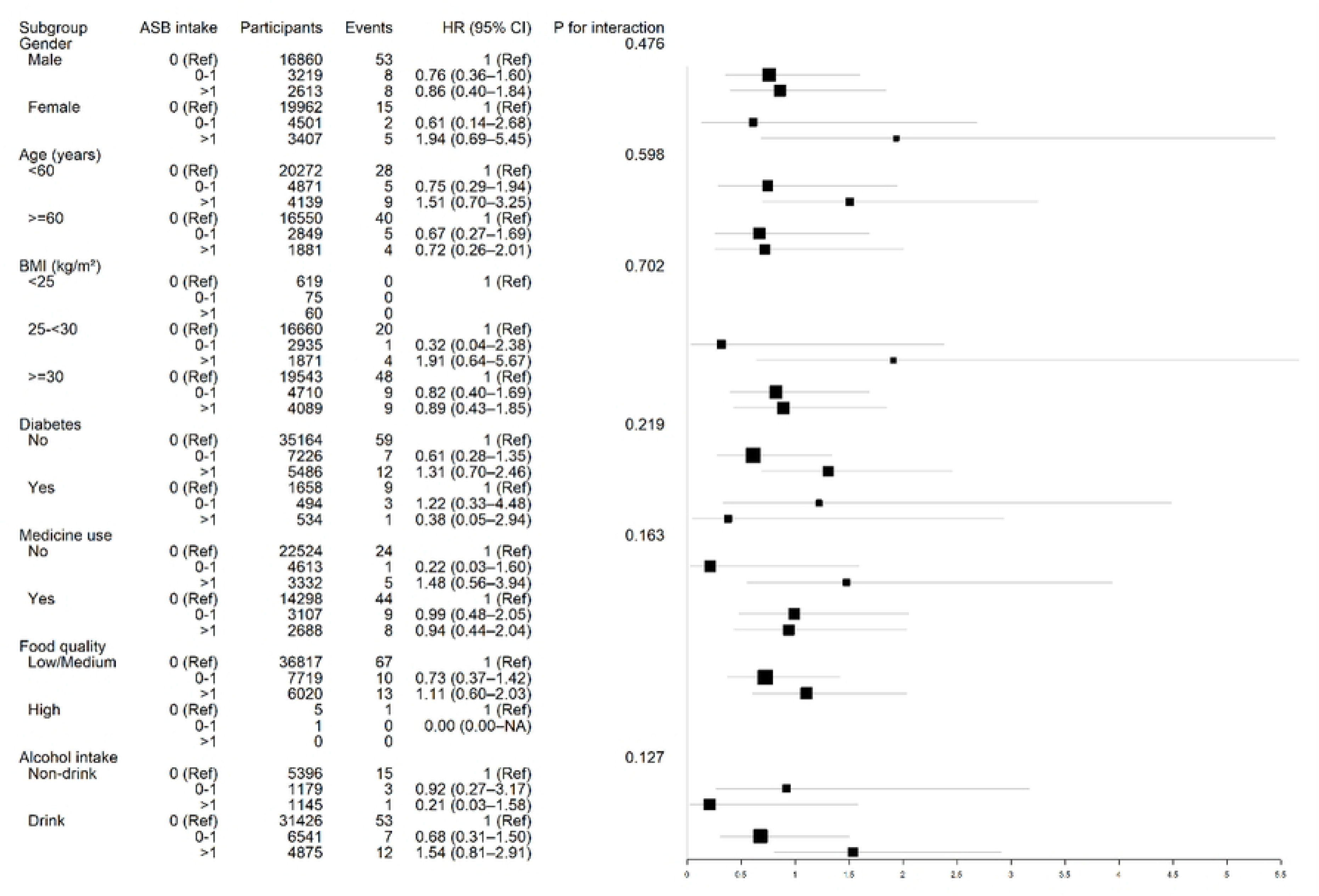
Subgroup analyses for the association between daily ASBs intake and liver-related adverse events/ liver-related death

## References

1. Zheng Y, Hou J, Guo S, Song J. The association between the dietary index for gut microbiota and metabolic dysfunction-associated fatty liver disease: a cross-sectional study. Diabetol Metab Syndr. 2025;17: 17. doi:10.1186/s13098-025-01589-9

2. Agyapong G, Dashti F, Banini BA. Nonalcoholic liver disease: epidemiology, risk factors, natural history, and management strategies. Ann N Y Acad Sci. 2023;1526: 16–29. doi:10.1111/nyas.15012

3. Wang L, Wang H, Wu J, Ji C, Wang Y, Gu M, et al. Gut microbiota and metabolomics in metabolic dysfunction-associated fatty liver disease: interaction, mechanism, and therapeutic value. Front Cell Infect Microbiol. 2025;15: 1635638. doi:10.3389/fcimb.2025.1635638

4. Yu Z, Chen M, Gu S, Wang C, Feng P, Lin G. Sugar-sweetened beverage consumption predicts metabolic associated fatty liver disease in patients with type 2 diabetes mellitus. Front Endocrinol. 2025;16: 1651370. doi:10.3389/fendo.2025.1651370

5. Ye B, Liu Y, Zhang J, Gong J, Zhu B, Chen W. Sweetened beverage intake and risk of incident kidney stone: results from the UK biobank. Front Nutr. 2026;13: 1844118. doi:10.3389/fnut.2026.1844118

6. Beigrezaei S, Raeisi-Dehkordi H, Hernández Vargas JA, Amiri M, Artola Arita V, van der Schouw YT, et al. Non-sugar-sweetened beverages and risk of chronic diseases: an umbrella review of meta-analyses of prospective cohort studies. Nutr Rev. 2025;83: 663–674. doi:10.1093/nutrit/nuae135

7. Kotha A, Sanyal AJ, Tariq R. Precision medicine in inflammatory bowel disease: the emerging role of metabolic dysfunction. J Pers Med. 2026;16: 139. doi:10.3390/jpm16030139

8. Sudlow C, Gallacher J, Allen N, Beral V, Burton P, Danesh J, et al. UK biobank: an open access resource for identifying the causes of a wide range of complex diseases of middle and old age. PLoS Med. 2015;12: e1001779. doi:10.1371/journal.pmed.1001779

9. Bazan A, Plichta O, Radzinska N. Metabolic dysfunction-associated steatotic liver disease: current diagnostic pathways, noninvasive fibrosis assessment, and therapeutic strategies. Cureus. 2026;18: e108965. doi:10.7759/cureus.108965

10. López-González ÁA, Tárraga López PJ, Piña Dabreu MS, Rodas Cañellas L, Busquets-Cortés C, Ramírez-Manent JI. Waist-to-height ratio as a simple anthropometric marker for identifying individuals at high risk of MASLD: a large population-based analysis using the fatty liver index. Metabolites. 2026;16: 246. doi:10.3390/metabo16040246

11. Lim TS, Chung SJ, Jeon J, Kim JK, Kim J. The influence of metabolic dysfunction-associated steatotic liver disease and body mass index on the incidence of alzheimer disease: a nationwide cohort study. Gut Liver. 2026;20: 107–116. doi:10.5009/gnl250079

12. Pan X, Lv J, Liu M, Li Y, Zhang Y, Zhang R, et al. Chronic systemic inflammation predicts long-term mortality among patients with fatty liver disease: data from the national health and nutrition examination survey 2007-2018. PLOS One. 2024;19: e0312877. doi:10.1371/journal.pone.0312877

13. Yang S, Mo R, Wang W, Zhen P, Han W. Development of a machine learning-based model for prediction of diabetes risk in patients with metabolic dysfunction-associated steatotic liver disease (MASLD). BMJ OPEN. 2026;16: e107239. doi:10.1136/bmjopen-2025-107239

14. Baek S-U, Kim T, Lee Y-M, Won J-U, Yoon J-H. Association between dietary quality and non-alcoholic fatty liver disease in korean adults: a nationwide, population-based study using the korean healthy eating index (2013-2021). Nutrients. 2024;16: 1516. doi:10.3390/nu16101516

15. Hall RL, George ES, Tierney AC, Reddy AJ. Effect of Dietary Intervention, with or without Cointerventions, on Inflammatory Markers in Patients with Nonalcoholic Fatty Liver Disease: A Systematic Review and Meta-Analysis. Advances in Nutrition. 2023;14: 475–499. doi:10.1016/j.advnut.2023.01.001

16. Espinosa A, Pacheco LS, Wan Y, Sun Q, Hu FB, Tobias DK, et al. Artificially sweetened beverages and weight change: findings from 3 prospective cohort studies of United States adults. Am J Clin Nutr. 2026;123: 101261. doi:10.1016/j.ajcnut.2026.101261

17. Graneri LT, Mamo JCL, D’Alonzo Z, Lam V, Takechi R. Chronic intake of energy drinks and their sugar free substitution similarly promotes metabolic syndrome. Nutrients. 2021;13: 1202. doi:10.3390/nu13041202

18. Alawadi AA, Vijay A, Grove JI, Taylor MA, Aithal GP. The development of a food-group, tree classification method and its use in exploring dietary associations with metabolic dysfunction-associated steatotic liver disease (MASLD) and other health-related outcomes in a UK population. Metab OPEN. 2025;25: 100351. doi:10.1016/j.metop.2025.100351

19. Sun T, Yang J, Lei F, Huang X, Liu W, Zhang X, et al. Artificial sweeteners and risk of incident cardiovascular disease and mortality: evidence from UK Biobank. Cardiovasc Diabetol. 2024;23: 233. doi:10.1186/s12933-024-02333-9

20. Lin Z, Zeng M, Ji Y, Wang L, Wu Y, Zhang T, et al. Association of sugar-sweetened, artificially sweetened, and unsweetened tea with all-cause and cause-specific mortality: a prospective cohort study in UK biobank. BMC Public Health. 2025;25: 2834. doi:10.1186/s12889-025-24063-7

21. Kim J-H, Kwon Y-J, Lee Y, Han T, Lim MY, Heo S-J, et al. Associations of Individual Beverage Types and Substitution with Dementia Risk: A UK Biobank Cohort Study. The Journal of nutrition, health and aging. 2026;30: 100740. doi:10.1016/j.jnha.2025.100740

22. Dan L, Fu T, Sun Y, Ruan X, Lu S, Chen J, et al. Associations of sugar-sweetened beverages, artificially sweetened beverages, and natural juices with cardiovascular disease and all-cause mortality in individuals with inflammatory bowel disease in a prospective cohort study. Therap Adv Gastroenterol. 2023;16: 17562848231207305. doi:10.1177/17562848231207305

23. Ayoub-Charette S, Nguyen M, Chiavaroli L, Mattes RD, de Souza RJ, Khan TA, et al. Low- and no-calorie sweeteners and health: unravelling the evidence and controversy. Appl Physiol Nutr Metab = Physiol Appl Nutr Metab. 2026;51: 1–11. doi:10.1139/apnm-2025-0440

24. Ayoub-Charette S, Kavanagh ME, Khan TA, Sievenpiper JL. Reconciling conflicting evidence on low- and no-calorie sweeteners and cardiometabolic outcomes: an umbrella review using naïve and bias-adjusted methods. Appl Physiol Nutr Metab = Physiol Appl Nutr Metab. 2025;50: 1–26. doi:10.1139/apnm-2025-0068

25. Khattab R. Artificial sweeteners and gut microbiota: mechanistic insights and implications for metabolic health. Curr Nutr Rep. 2026;15: 47. doi:10.1007/s13668-026-00768-y

26. Tan HS, Gilcharan Singh HK, Mariappan V, Jamil NA, Mustapa SN, Misra S. Short-term effects of nonnutritive sweetener (sucralose and saccharin) consumption on glycemic control and gut microbiota in patients with type 2 diabetes: protocol for a double-blind, randomized, placebo-controlled, crossover trial. JMIR Res Protoc. 2025;14: e82695. doi:10.2196/82695

27. Zhao L, Zhang X, Coday M, Garcia DO, Li X, Mossavar-Rahmani Y, et al. Sugar-sweetened and artificially sweetened beverages and risk of liver cancer and chronic liver disease mortality. JAMA. 2023;330: 537–546. doi:10.1001/jama.2023.12618

28. Watling CZ, Zhao L, Zhang X, Deubler E, Gonzalez-Feliciano AG, Graubard BI, et al. Artificially sweetened and sugar-sweetened beverage intake and risk of liver cancer. JAMA Netw Open. 2026;9: e2617754. doi:10.1001/jamanetworkopen.2026.17754

29. Zhang Y, Xiao W, Zhu X, Zhu Y, Yang Z, Ma J. Association of dietary nutrient intake with reproductive lifespan in women: a cross-sectional analysis using weighted quantile sum regression in NHANES 2001-2018. BMC Women’s HEALTH. 2025;26: 17. doi:10.1186/s12905-025-04224-x

30. Yang L, Shen X, Seyiti Z, Tang J, Lu J, Kasimujiang A, et al. Dietary niacin intake and mortality outcomes in hypertensive populations: analysis from NHANES 2003-2016. J Health Popul Nutr. 2025;44: 206. doi:10.1186/s41043-025-00976-2

31. Bragança MLBM, Bogea EG, de Almeida Fonseca Viola PC, Dos Santos Vaz J, Confortin SC, Menezes AMB, et al. High consumption of sugar-sweetened beverages is associated with low bone mineral density in young people: the brazilian birth cohort consortium. Nutrients. 2023;15: 324. doi:10.3390/nu15020324

32. Yu Z, Ley SH, Sun Q, Hu FB, Malik VS. Cross-sectional association between sugar-sweetened beverage intake and cardiometabolic biomarkers in US women. Br J Nutr. 2018;119: 570–580. doi:10.1017/S0007114517003841

33. Rousham EK, Goudet S, Markey O, Griffiths P, Boxer B, Carroll C, et al. Unhealthy food and beverage consumption in children and risk of overweight and obesity: a systematic review and meta-analysis. Adv Nutr. 2022;13: 1669–1696. doi:10.1093/advances/nmac032

34. do Prado CB, Cattafesta M, Martins CA, Pedraza DF, Silva YFR, Ferreira JRS, et al. The cutoff points of body roundness index for predicting metabolic syndrome in the brazilian population among 18-59 years. Sci Rep. 2025;15: 13084. doi:10.1038/s41598-025-97212-y

